# Predicting risk of pancreatic cancer in individuals with new-onset type-2 diabetes in primary care: protocol for the development and validation of a clinical prediction model (QPancreasD)

**DOI:** 10.1101/2021.12.22.21268161

**Authors:** Pui San Tan, Ashley Kieran Clift, Weiqi Liao, Martina Patone, Carol Coupland, Rachael Bashford-Rogers, Shivan Sivakumar, David Clifton, Stephen P Pereira, Julia Hippisley-Cox

**Author notes:** Corresponding author: Professor Julia Hippisley-Cox, Nuffield Department of Primary Care Health Sciences, University of Oxford Radcliffe Primary Care Building, Radcliffe Observatory Quarter, Woodstock Road, Oxford OX2 6GG, UK.

## Abstract

**Background:** Pancreatic cancer continues to have an extremely poor prognosis in part due to late diagnosis. 25% of pancreatic cancer patients have a prior diagnosis of diabetes, and hence identifying individuals at risk of pancreatic cancer in those with recently diagnosed type 2 diabetes may be a useful opportunity to identify candidates for screening and early detection. In this study, we will comparatively evaluate regression and machine learning-based clinical prediction models for estimating individual risk of developing pancreatic cancer two years after type 2 diabetes diagnosis.

**Methods:** In the development dataset, we will include adults aged 30-84 years with incident type-2 diabetes registered with QResearch primary care database. Patients will be followed up from type-2 diabetes diagnosis to first diagnosis of pancreatic cancer as recorded in any one of primary care records, hospital episode statistics, cancer registry data, or death records. Cox-proportional hazards models will be used to develop a risk prediction model for estimating individual risk of developing pancreatic cancer during up to 2 years of follow-up. We will perform variable selection using a combination of clinical and statistical significance approach i.e. HR <0.9 or >1.1 and p<0.01. Linear predictors and baseline survivor function at 2 years will be used to compute absolute risk predictions.

Internal-external cross-validation (IECV) framework across geographical regions within England will be used to assess performance and pooled using random effects meta-analysis using: (i) model fit in terms of variation explained by the model Royston & Sauerbrei’s R2D, (ii) calibration slope and calibration-in-the-large, and (iii) discrimination measured in terms of Harrell’s C and Royston & Sauerbrei’s D-statistic.

Further, we will evaluate machine learning (ML) approaches for the clinical prediction model using neural networks (NN) and XGBoost. The model predictors and performance of these will be compared with the results of those derived from the regression-based strategy.

**Discussion:** The proposed study will develop and validate a novel risk prediction model to aid early diagnosis of pancreatic cancer in patients with new-onset diabetes in primary care. With an enhanced decision-risk tool for use at point-of care by general practitioners to assess pancreatic cancer risk, it may improve decision-making so that at-risk patients are rapidly prioritised to aid early diagnosis of pancreatic cancer in patients with newly diagnosed diabetes.

## Background

Pancreatic cancer and its predominant subtype pancreatic ductal adenocarcinoma^1^ remain hard-to-treat and patients’ prognoses are extremely poor, with only 25.4% of patients surviving beyond one year after diagnosis.^2^ Poor prognosis is driven by typically late presentation of advanced disease where surgical treatment is not feasible. In the absence of a screening programme, symptoms-based detection are complicated by the often asymptomatic nature or vague symptoms at early stage disease.^3^ Considering that, integration of symptom information and other factors may be informative to identify higher risk individuals suitable for higher-risk referral pathways, or recruitment into screening or prevention trials.

Type 2 diabetes is a common condition with up to 7% of the UK population affected.^4^ The association between type 2 diabetes and pancreatic cancer^3^ could be due to either type 2 diabetes driving pancreatic cancer pathogenesis, or type 2 diabetes starting due to local tumour effects disrupting the endocrine function of the pancreas.^5^ Regardless, as approximately 1 in 4 people with pancreatic cancer have diabetes,^6^ focussing on people recently diagnosed with type 2 diabetes may be a useful opportunity to identify enriched sub-populations in terms of pancreatic cancer risk, in which screening and early detection strategies could be targeted in primary care.^7-10^

Earlier studies have shown that diabetes was associated with the risk of pancreatic cancer with a 1.8-2.2 fold increase, compared to patients without diabetes, and this association was strongest when diabetes onset was less than 2 years prior to the diagnosis of pancreatic cancer. ^7,8,11,12^ Further studies also suggested that particular characteristics of diabetes control could be associated with risk of pancreatic cancer; including weight loss and unstable glucose control.^8,13,14^ In addition, some studies have also suggested that glucose lowering therapies such as insulin, sulphonylureas, and DPP4-inhibitors are associated with the risk of pancreatic cancer.^10^ A recent QResearch case-control study including over 24,000 incident pancreatic cancer cases from 2000-2019 identified a set of ‘red flag’ symptoms and medical conditions that were associated with increased risk of being diagnosed with pancreatic cancer. Symptoms reported included haematemesis, jaundice and dysphagia in the last 3 months prior to pancreatic cancer diagnosis while risk factors included smoking, chronic pancreatitis, prior cancers, and type 2 diabetes mellitus (T2D).^6^ In the study, 24% of patients with pancreatic cancer had a prior diagnosis of type 2 diabetes, compared to 12.5% in people without pancreatic cancer, which also found 15 additional symptoms and 6 risk factors compared with the existing QCancer tool.^11,12^

In 2015, NICE updated the suspected cancer pathway referral guidelines in which patients who are aged 60 and over and presenting with both weight loss and new-onset diabetes are to be given urgent two-week referral direct access CT scans or an ultrasound scan if CT is not available.^15^ However, it remains unknown to what extent this guidance has been implemented by general practitioners in primary care or whether the threshold of age 60 is appropriate and if a lower age threshold might improve ascertainment rates.

Approximately 1 in 100 of new-onset type 2 diabetes patients have pancreatic cancer within three years.^16^ and it remains impractical to refer everyone with type 2 diabetes for pancreatic cancer scans.^15^ Hence, a multivariable risk stratification tool for adults with type 2 diabetes could inform prioritisation of higher-risk patients. A US study developed and validated a risk prediction model for developing pancreatic cancer in a relatively small sample of 1,561 new-onset diabetes patients with 16 (1%) of patients developing pancreatic cancer within three years.^17^ This study stratified patients into high, intermediate and low risk for pancreatic cancer. However this study has two main limitations, firstly the model was developed using univariable screening of predictors, which had not taken into account the effect of other confounder variables and may wrongly rule out potentially useful predictors.^18^ Secondly, the model was developed using a relatively small sample size with no evaluation of overfitting,^19^ as well as the lack of reporting of calibration and discrimination measures. Another Taiwan study also developed a risk prediction model for pancreatic cancer in patients with new-onset diabetes.^20^ However, this study had used a binary outcome classification of pancreatic cancer, had not evaluated risk prognosis in a time-to-event manner, and did not evaluate model performance using calibration and discrimination measures. Hence, taken together, risk models in current literature suggests further need for more robustly evaluated risk prediction model.

This study proposes to develop and validate clinical prediction models to estimate short term risk of pancreatic cancer diagnosis in the two years immediately following a type 2 diabetes diagnosis, and improve on existing tools which are already used by GPs to identify those at increased risk of having an undiagnosed pancreatic cancer.^11,12,21^ Existing tools have been developed using established statistical approaches, however machine learning approaches may offer additional insights and have improved performance. Despite growing interest in the possibility of using machine learning approaches for prediction modelling,^22^ robust, fair and transparently reported comparisons between machine learning and regression-based approaches are not yet common in the literature.

In this study, we aim to comparatively evaluate regression and machine learning-based clinical prediction models for estimating individual risk of having an as yet undiagnosed pancreatic cancer after type 2 diabetes diagnosis.

## Methods

### Study design

We will perform a cohort study of adults aged 30-84 years registered with QResearch practices for at least a year beginning 01 Jan 2010 to last study update. We will identify patients with newly diagnosed type 2 diabetes who will enter the cohort on the earliest date of a recorded type 2 diabetes diagnosis. Patients will be followed up from date of type 2 diabetes diagnosis until the earliest of the date of diagnosis of pancreatic cancer, or censoring due to death, loss to follow-up, or end of 2 years from the date of type 2 diabetes diagnosis.

### Data sources

QResearch primary care database (version 46) with individual-level linkages to the national cancer registry (Public Health England), Hospital Episode Statistics (HES), and Office for National Statistics death registry in England.

### Study population

#### Inclusion criteria

Adults aged 30-84 years with incident type-2 diabetes during the study period, who have been registered with QResearch for at least a year. Incident type 2 diabetes will be defined as any individual with a first record of type 2 diabetes in the GP records during the study period.

### Exclusion criteria

Patients with an existing diagnosis of pancreatic cancer prior to the diagnosis of type 2 diabetes. Further we will also exclude those with records of diabetes medication use prior to type 2 diabetes diagnosis as they could be suggestive of prevalent diabetes.

### Outcome

The outcome of the study will be the first diagnosis of pancreatic cancer, as recorded in any one of: primary care records using SNOMED/Read codes, hospital episode statistics, cancer registry data, or Office for National Statistics death records using ICD10 codes as published https://www.qresearch.org/qcode-group-library/

### Candidate predictor variables

We will include demographics, lifestyle factors, comorbidities, symptoms, laboratory tests and drugs as below as informed by previous pancreatic cancer risk models and prognostic studies in the literature.

**Demographics** – age at index date (first diagnosis of diabetes), sex, Townsend deprivation quintile, ethnicity (up to 9 categories)

**Lifestyle** – alcohol intake, smoking status, body mass index (BMI)^6^

### Conditions (prior to T2D diagnosis)

Acute pancreatitis^6^

Chronic pancreatitis^6^

Venous thromboembolism^6^

Cushing’s syndrome^6^

Pancreatic cyst^6^,

Family history of gastro-intestinal cancer^6^

Asthma^23,24^,

Hepatitis C^25^

Peptic ulcer disease^26^,

Helicobacter pylori infection^27^

Gastro-oesophageal reflux^28^

Cystic fibrosis^29^

Cholangitis^6^

Gall bladder problems^30^,

Family history of diabetes^31^,

Hypertension^32^,

Prior breast cancer^24^

Prior ovarian cancer^24^

Prior prostate cancer^24^

Coeliac disease^24^

HIV/AIDS^24^

### Symptoms (on/within 6 months prior to T2D diagnosis) (n=23) ^6,11,12,21^

Haematemesis, jaundice, dysphagia, diarrhoea, change in bowel habit, vomiting, indigestion, abdominal mass or lump, abdominal pain, weight loss, constipation, steatorrhea, abdominal distension, nausea, flatulence, heartburn, fever, tiredness fatigue and sleepiness, loss of appetite, itching, back pain, excessive thirst, dark urine

### Tests/measurements (on/within 6 months prior to T2D diagnosis)

Glucose test - HbA1C^6 21^

Liver function tests^33^ –ALT, bilirubin

Creatinine^34^

Blood counts^34^- haemoglobin (anaemia), white blood cell, platelet

Inflammatory markers - ESR^34^, CRP^35 34^

### Drugs (current users with at least 3 prescriptions on/within 12 months prior to T2D diagnosis)

Calcium channel blockers^32^

Proton-pump inhibitors (PPI)^36,37^

H2-receptor blockers^37^

Bisphosphonates^38^

Aspirin^39^

Statins^40^

Digoxin^41^

### Sample size considerations

We estimated the minimum required sample size required using pmsampsize in Stata.^42^ Assuming an estimated (conservative) rate of 0.3% diagnosis of pancreatic cancer within 2 years of diabetes diagnosis,^16^ up to 100 predictor parameters being considered (see variable list above, categorical variables are counted as number of categories-1), R^2^ (0.0105; 15% of max Cox-Snell R-squared 0.07), 2 years follow-up, and mean follow-up of 2 years, we estimate that we would need 85,214 persons with type 2 diabetes (events per predictor parameter =5.11). Currently, we expect to have over 300,000 incident type 2 diabetes cases in QResearch during the study period, so we can be confident that the study will be sufficiently powered. We acknowledge that at present there are no established approaches to calculating adequate minimal sample size for machine learning prediction models.^43^ Simulation studies suggest that the minimal sample size may need to be higher than for regression models, however such studies are restricted to binary outcome prediction.^43^

### Handling of missing data

Where data is missing for ethnicity, deprivation, BMI, alcohol intake, and smoking, missing values will be imputed using multiple imputation with chained equations using all outcome and exposure variables in the model (including their transformations where relevant). We will also include Nelson– Aalen estimator of the cumulative baseline hazard in the imputation model. Ten imputations will be generated and Rubin’s rules^44^ will be used to pool model coefficients and their standard errors across the imputed datasets. We will assess the extent and pattern of missingness in laboratory test variables e.g. if missingness is more than 50%, we will explore patterns of missingness to ascertain whether multiple imputation under the missing at random assumption is valid, if it is these variables will be included in the multiple imputation model. For the machine learning modelling approaches, as these models do not have parameters and standard errors in a classical sense, we will use a stacked imputation approach for model fitting, with model evaluation using imputed datasets (performance metrics calculated in each imputation and combined using Rubin’s rules).

### Statistical methods

#### Descriptive analysis

We will present descriptive analyses of the characteristics of the cohort, including the prevalence of baseline characteristics. We will also report on the proportion of type 2 diabetes patients who were referred for a CT scan and the proportion referred for an abdominal ultrasound within a month of diagnosis of type 2 diabetes who also had evidence of weight loss (in line with NICE guidance) ^15^, stratified by age (under 60; 60+), by sex, region and ethnicity. We will compare this over time, focusing on the period before and after the 2015 guidance. We will also report on the route to diagnosis e.g. 2 week-wait referral for patients with pancreatic cancer in the same period.

#### Model development

We will build the prognostic equations for time to developing pancreatic cancer from the date of diagnosis of type 2 diabetes. This will use time to event models e.g. Cox-proportional hazards model to model predictor variables listed above with robust variance estimates to account for clustering within general practices. Non-linear relationships of continuous variables with outcome will be considered and modelled with fractional polynomials. In the case of non-proportional hazards, we explore time-varying risks using flexible parametric models^45^ to explore potential changing risks over time of developing pancreatic cancer from type 2 diabetes diagnosis.

#### Variable selection

We will fit the model using a combination of clinical and statistical significance approach. Binary and categorical variables will be kept in the model if they show clinical significance i.e. HR <0.9 or >1.1 and statistical significance (p<0.01). A p-level of 0.01 is used to ensure a stringent criterion is applied to keep only the most useful predictor variables. For continuous predictors, we will model these using fractional polynomial (FP) terms to account for non-linear associations and include based on statistical significance (p<0.01). Further, established risk factors based on prior knowledge (e.g. age, sex) will be retained in the model irrespective of clinical or statistical significance. Pre-specified interactions between relevant variables will also be examined, for example between age and family history of gastrointestinal cancer.

#### Formulation of risk equations

We will combine linear predictors with the baseline survivor function at 2 years to compute absolute risk predictions using the formula:

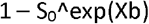

*Where S_0_ is the baseline survivor function at 2 years; Xb is the individual linear predictor*.

The baseline survivor function will be estimated by centring all continuous variables at their mean values and setting binary variables at zero.

#### Model evaluation and validation

We will use an internal-external cross-validation (IECV) framework across geographical regions within England to assess performance and quantify performance heterogeneity of a model fitted to the entirety of the available data.^1,2^ In the IECV framework (See Figure 1), the dataset will be split by the individual geographical regions in England. Individual regions will be iteratively ‘held out’ as a test set, and used to validate a model fitted to all data from remaining regions. This will be repeated so that all regions sequentially contribute to model fitting and evaluation, and a linear predictor is generated for each individual when they are in the held-out set. The following performance metrics will be calculated at a regional level and pooled using random effects meta-analysis: (i) model fit in terms of variation explained by the model (Royston & Sauerbrei’s R^2^ _D_)^46^, (ii) calibration slope and calibration-in-the-large, and (iii) discrimination measured in terms of Harrell’s C^47^ and Royston & Sauerbrei’s D-statistic^48^. Using the individual predictions generated from IECV, calibration plots will be generated for each held-out region comparing mean predicted risks and observed risks (derived from Kaplan-Meier estimates) at 2 years by tenths or twentieths of predicted risks. The sensitivity and specificity of the model will be assessed at the top 1%, 5% and 10% of predicted risks. In addition, we will perform sensitivity analyses to compare robustness of results between complete-case and imputed analyses.

**Figure 1:**
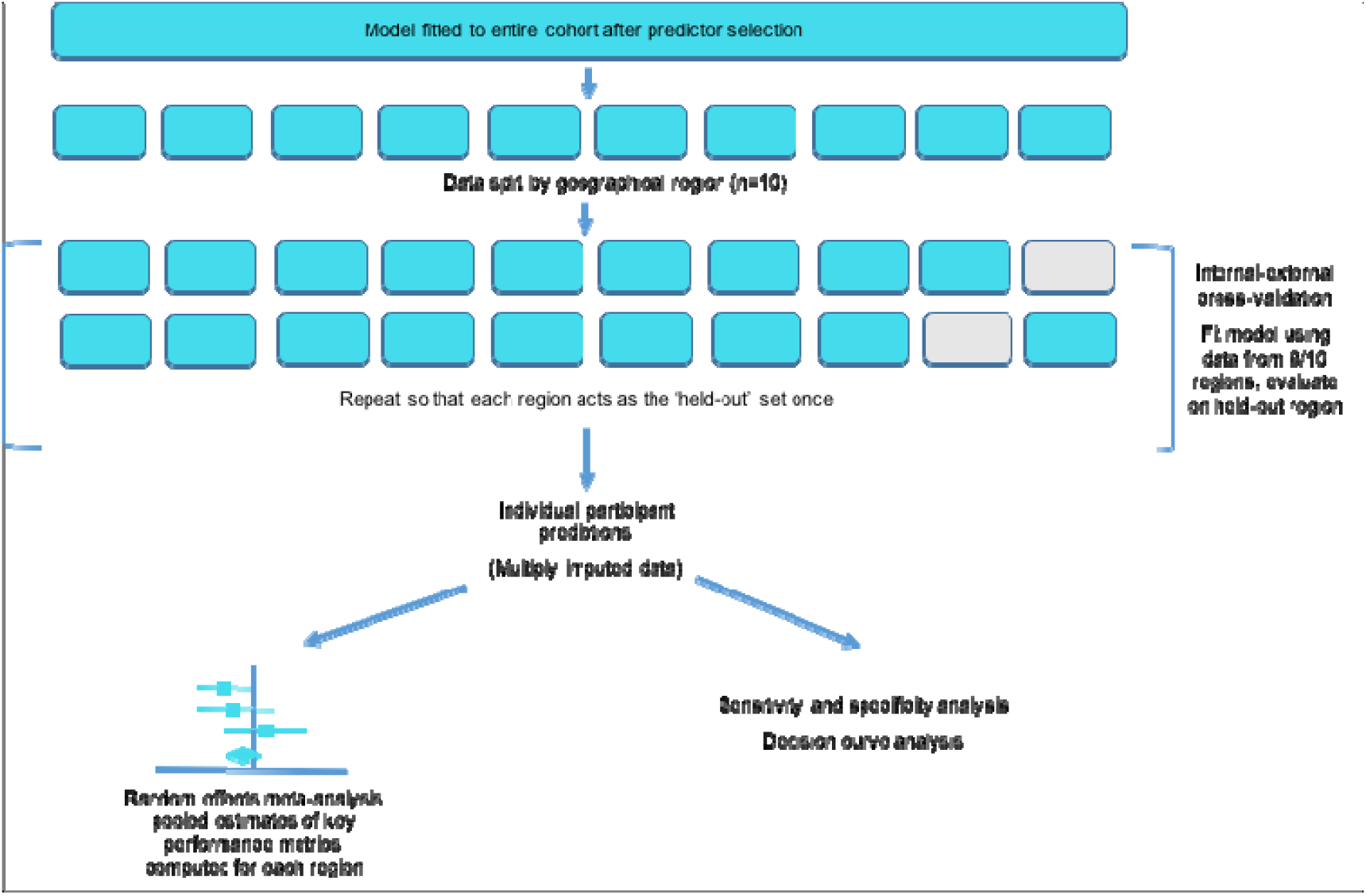
Schematic diagram of internal-external cross validation (IECV)

Further, we will explore performance heterogeneity by clinically relevant sub-group, e.g. ethnicity. We will also explore the use of smoothed calibration plots using the pseudo-values based approach of Royston to explore calibration across the entire spectrum of predicted risks.^49-53^

### Machine learning

We will explore machine learning (ML) approaches for the clinical prediction model, and compare the results of these to those derived from the regression-based model. Two forms of ML model will be explored: neural networks (NN) and XGBoost, which will be adapted to handle time-to-event data. We will explore three methods of predictor parameter selection: ML1) Include the variables selected in the regression modelling approach, ML2) A full model with all candidate predictors to estimate a ‘benchmark’ for a maximally complex model, and ML3) variable selection based on a threshold of relative variable importance after fitting the previous ‘maximally complex’ models.

Variants of neural networks exist which seek to handle survival data, such as DeepSurv ^54^ (which uses a Cox partial likelihood loss function and can output a non-linear predictor), and DeepHit^55^ (adapted for competing risks). Furthermore, a variant of XGBoost^56^ allows a boosted tree-based approach to minimise the Cox partial likelihood and outputs a ‘hazard ratio’, i.e. exponentiated (non)linear predictor. Other approaches include the use of jack-knife pseudo-values for the survival function or cumulative incidence function, which were initially developed for a regression setting, ^49-53^ but have been adapted for use in ML models such as neural networks or random forests.^57-60^ These use the derived pseudo-values as a continuous target variable (rather than separate time and status variables), relax the proportional hazards assumption, and can enable direct modelling of event probabilities without the need to separately estimate baseline functions for combination with a model output. Therefore, here we will utilise the pseudovalues-based approach.

Jack-knife pseudo-values of the Kaplan-Meier failure function at 2 years follow-up will be estimated for the entire cohort using the ‘stpsurv’ command in Stata. These will be used as continuous outcome variables to fit a neural network model and an XGBoost model to the entire cohort data, which will then undergo evaluation with IECV. For the NN, continuous variables will be min-max scaled (to between 0-1), and for both models, categorical variables with more than 2 levels will be converted to dummies. To handle missing data, the imputations obtained earlier will be stacked to form a single ‘long’ dataset, with each observation assigned a weight of 1/number of imputations. This enables all imputations to be used in the model fitting by weighting the contribution of each individual’s ‘variant’ to the loss function. This ‘stacked and weighted’ approach circumvents the issue that these models cannot pool parameters/standard errors in the same fashion as a regression model. The neural network will have a single output node with a linear activation function, ReLU activation functions on nodes in hidden layers, use the Adam optimiser, and use the root mean squared error (RMSE) between predicted and observed pseudo-values as a loss function. The XGBoost model will have a regression objective, with the same loss function as the NN.

Ten-fold cross-validation will be used to identify optimal model hyperparameters to minimise model root mean squared error (RMSE) (Table 1). Optimising the RMSE avoids purely focussing on maximising a discrimination metric (such as Harrell’s C) with disregard to model calibration. The optimal configurations will be identified using Bayesian Optimisation, ^61^ and the final model fitted to the entire cohort using these. The performance of these final models will be estimated using IECV with the data split by region (as for the regression modelling) - within this IECV framework, hyperparameter tuning will be repeated with 10-fold cross-validation in each cycle, thus constituting a form of ‘nested’ cross-validation ^62^ and avoiding the inherent bias in using the same dataset for simultaneous tuning and evaluation. Individual level predictions will be generated when participants are in the iteratively held-out set, and analysed in the same way as the regression models, including the estimation of key metrics in each imputed dataset and pooling in accordance with Rubin’s rules.

**Table 1:** Model hyperparameters and tuning ranges

| Model type | Hyperparameter | Range explored in tuning |
| --- | --- | --- |
| Neural network<br>*Multilayer perceptron, fully connected layers, ReLU activation functions in hidden layers, linear activation in single node output layer, Adam optimiser, batch size 128* | Initial learning rate | 0.0001 to 0.1 |
|  | Number of hidden layers | 1 to 20 |
|  | Number of nodes in hidden layers | [number of inputs to 2x] |
|  | L2 regularisation | 0.0001 to 0.5 |
|  | Number of training epochs | 1 to 100 |
| XGBoost<br>*Regression-objective booster,* | Number of boosting rounds | 1 to 1000 |
|  | Maximum tree depth | 1 to 10 |
|  | Learning rate (eta) | 0.001 to 0.1 |
|  | Subsampling ratio | 0.2 to 0.8 |
|  | Column sampling ratio per tree | 0.1 to 0.8 |
|  | Column sampling ratio per node | 0.1 to 0.8 |
|  | Lambda | 0 to 1000 |

### Model comparisons

In addition, we will compare performance of the following models/interventions in terms of calibration, discrimination and decision curve analysis (where applicable).^63^ Decision curve analysis will be performed to evaluate clinical benefit of following interventions across different risk threshold probabilities compared to “screen all” and “screen none” approaches:

i. risk equation using regression approach
ii. risk equation using machine-learning approach (ANN) (variable selection: best performing as determined in ML1-3 above)
iii. risk equation using machine-learning approach (XGBoost) (variable selection: best performing as determined in ML1-3 above)
iv. existing risk equation (QCancer) ^11,12^
v. simple decision support risk equation using NICE guidelines; aged 60 and over presenting with both weight loss and new-onset diabetes^15^ (comparing sensitivity and specificity - this potentially could be implemented easily as opposed to more complex models above).

Statistical analyses will be performed using STATA (version 17)^64^ and/or R statistical software^65^ with results reported in accordance to TRIPOD guidelines.^66^

## Discussion

This study will develop and validate a novel risk prediction equation to aid early diagnosis of pancreatic cancer in patients with new-onset diabetes in primary care. An enhanced decision-risk tool for use at point-of care by GPs to assess pancreatic cancer risk, could improve decision-making so that most at-risk patients are rapidly prioritised to aid early diagnosis of pancreatic cancer amongst new-onset diabetes patients. We have the support of the PCUK Research Involvement Network lay members for this approach and, by diagnosing pancreatic cancer earlier, expect that this will improve the outcomes of pancreatic cancer once implemented in clinical practice.

Limitations of this study may include the possibility of bias due to missing data, i.e. information bias, which we will perform sensitivity analyses to compare robustness of results between complete-case and imputed analyses. There could also be potential ascertainment bias where pancreatic cancer cases might be misclassified – hence in this study we have taken a comprehensive approach, which is to capture pancreatic cancer cases from any of primary care records, hospital episode statistics, cancer registry data, or Office for National Statistics death records. Lastly, we also were not able to perform sample size calculations for the machine learning models as there is currently no established method to do so,^43^ however, we will use all data that is available in the dataset to develop the model and we expect that not all 100 predictor parameters will be included in the final model after variable selection.

In summary, this study will comparatively evaluate regression and machine learning-based clinical prediction models for estimating individual risk of developing pancreatic cancer after type 2 diabetes diagnosis. This risk model may have the potential to be implemented as a software tool in the NHS once it has been validated and subject to MHRA regulations and funding. If implemented, it may bring about a step-change in how patients are risk-assessed for pancreatic cancer in primary care.

## Data Availability

To guarantee the confidentiality of personal and health information, only the authors will have access to the data during the study in accordance with the relevant license and data sharing agreements. Access to QResearch data is according to the information on the QResearch website https://www.qresearch.org

## Ethical approval

This study has been independently peer-reviewed and received ethics approval by the QResearch Scientific board (18/EM/0400; project reference OX153).

## Patient and public involvement

We involved the participation of PCUK Research Involvement Network (RIN) lay members who have or cared for someone who have lived experience of pancreatic cancer to help with developing our research questions, protocol, and writing of lay summary.

## Funding

This project is funded by Pancreatic Cancer UK. AKC is supported by a Clinical Research Training Fellowship from Cancer Research UK (grant number C2195/A31310). JHC reports grants from National Institute for Health Research (NIHR), NIHR Biomedical Research Centre, Oxford, grants from John Fell Oxford University Press Research Fund, grants from Cancer Research UK (CR-UK) grant number C5255/A18085, through the Cancer Research UK Oxford Centre, grants from the Oxford Wellcome Institutional Strategic Support Fund (204826/Z/16/Z) and other research councils, during the conduct of the study. SPP is supported by the National Institute for Health Research University College London Hospitals Biomedical Research Centre.

## Contributor statement

JHC and PST obtained funding for the study. PST, AKC, JHC, and CC contributed to the design and methodological aspects of the study. AKC and JHC provided clinical input. PST, JHC and AKC contributed to the first draft. All authors contributed to its critical revision and approved the final version of the protocol.

## Competing interests

JHC is an unpaid director of QResearch, a not-for-profit organisation which is a partnership between the University of Oxford and EMIS Health who supply the QResearch database used for this work. JHC is a founder and shareholder of ClinRisk Ltd and was its medical director until 31st May 2019. ClinRisk Ltd produces open and closed source software to implement clinical risk algorithms (including the original development of pancreatic cancer prediction algorithms) into clinical computer systems. CC reports previous consultancy with ClinRisk Ltd outside the current work. AKC reports consulting fees from Mendelian, outside the scope of the current work. PST reports previous consultation with AstraZeneca and Duke-NUS outside the current work.

## Acknowledgements

We acknowledge the contribution of EMIS practices who contribute to QResearch and EMIS Health and the Universities of Nottingham and Oxford for expertise in establishing, developing, or supporting the QResearch database. This project involves data derived from patient level information collected by the NHS, as part of the care and support of cancer patients. The data are collated, maintained, and quality assured by the National Cancer Registration and Analysis Service, which is part of Public Health England (PHE). Access to the data was facilitated by the PHE Office for Data Release. The Hospital Episode Statistics data used in this analysis are reused by permission from NHS Digital, which retains the copyright in that data. We thank the Office for National Statistics (ONS) for providing the mortality data. NHS Digital, PHE, and the ONS bear no responsibility for the analysis or interpretation of the data. The views and opinions expressed by authors in this publication are those of the authors and do not necessarily reflect those of the UK National Institute for Health Research (NIHR) or the Department of Health and Social Care.

